# Screening for glaucoma with a novel eye movement perimetry technique based on continuous visual stimulus tracking

**DOI:** 10.1101/2025.10.17.25338104

**Authors:** A.C.L. Vrijling, M.J. de Boer, R.J. Renken, J.B.C. Marsman, J. Heutink, F.W. Cornelissen, N.M. Jansonius

**Affiliations:** Laboratory of Experimental Ophthalmology, University Medical Center Groningen, University of Groningen, Groningen, The Netherlands; Royal Dutch Visio, Centre of Expertise for Blind and Partially Sighted People, Huizen, The Netherlands; Center for Clinical Neuroscience and Cognition, University Medical Center Groningen, the Netherlands, University of Groningen, the Netherlands; Department of Clinical and Developmental Neuropsychology, University of Groningen, Groningen, The Netherlands; Department of Ophthalmology, University Medical Center Groningen, University of Groningen, Groningen, The Netherlands

**Keywords:** sensitivity and specificity, eye-tracking, continuous visual stimulus tracking, visual field, glaucoma, eye movements, screening performance

## Abstract

**Purpose:** Standard automated perimetry (SAP) is the gold standard for functional assessment in glaucoma. SAP can be too demanding for some groups of patients. Continuous visual stimulus tracking (SONDA: Standardized Oculomotor and Neurological Disorders Assessment) simplifies the perimetric task to following a moving stimulus on a screen. In this study we evaluated the screening performance of SONDA-based eye movement perimetry (SONDA-EMP) in glaucoma. To explore generalizability, we evaluated an experimental setup (SONDA-Eyelink) and a clinic-ready version (SONDA-Neon).

**Methods:** SONDA-Eyelink and SONDA-Neon measurements were performed in 100 cases with glaucoma (36, 36, and 28 with early, moderate, and severe glaucoma, respectively) and 100 age-similar controls. Participants monocularly tracked a moving stimulus (Goldmann size III) at 40% contrast (both setups) and 160% (SONDA-Eyelink). Eye movements were continuously recorded. Outcome was the agreement between gaze and stimulus position. We used previously collected glaucoma case-control data to build a continuous ‘glaucoma screening score’. This score was used for an ROC-analysis applied to the current, independently collected dataset. We predefined good screening performance as: at 95% specificity, a sensitivity of at least 50%, 90%, and 100% for early, moderate, and severe glaucoma, respectively.

**Results:** At 95% specificity, the sensitivity of SONDA-Eyelink was 58, 94, and 100% at 40% contrast and 56, 97, and 100% at 160% contrast for early, moderate, and severe glaucoma, respectively. Sensitivity was 53, 94, and 100% for SONDA-Neon.

**Conclusions:** SONDA-EMP is a novel, fast, and intuitive method to screen for visual function loss in glaucoma.

## Introduction

Glaucoma is a chronic, progressive disease that can lead to irreversible visual function loss, and ultimately, blindness. In its early stages, glaucoma is typically asymptomatic, making timely detection and diagnosis challenging. However, early detection and treatment are essential as disease progression can be significantly slowed with appropriate intervention.^1,2^ Visual field (VF) testing is important for detecting, characterizing, and monitoring vision loss in glaucoma. The current gold standard for VF assessment is standard automated perimetry (SAP). SAP presents stimuli of varying luminance at fixed locations across the VF in a systematic way, requiring the individual to maintain steady central fixation and press a button upon perceiving a stimulus. Performing SAP requires prolonged focused attention, understanding of the test, as well as multi-tasking. These requirements may hamper the use of SAP in children, elderly, and persons with cognitive and/or motor impairments, critically limiting the quality of ophthalmic and rehabilitation care. Given these limitations, there is a need for faster, more intuitive (that is, less cognitive demanding), and more accessible methods to assess visual function loss such as due to glaucoma.

Several studies have demonstrated the efficacy of eye movement perimetry (EMP) in identifying glaucomatous VF defects.^3–10^ EMP replaces the manual responses as used in SAP by eye-movement-based responses that can be recorded using an eye tracker.^11,12^ These responses can be quantified using the saccadic reaction time when the gaze moves from a fixation target to a stimulus presented somewhere in the VF.^4,5,7,8,10,12^ A next step in the development of EMP was the development of SONDA (Standardized Oculomotor and Neurological Disorders Assessment). SONDA uses a continuous visual stimulus tracking paradigm, which additionally eliminates the need to return to a fixation target after each saccade, and uses all sampled data acquired over the entire measurement session to assess the integrity of the visual system.^13^

In our previous studies using the SONDA-based eye movement perimetry technique (SONDA-EMP), we demonstrated that ‘tracking performance’ (defined as the agreement between gaze and stimulus position), improves with increasing stimulus contrast in healthy participants. We also found that, with ageing, a higher stimulus contrast is needed to maintain performance.^14^ Furthermore, we showed that SONDA-EMP can be used to detect and quantify visual function loss due to glaucoma^15^ as well as acquired brain injury (*Vrijling et al., submitted*). For the detection of glaucoma cases, we determined that a stimulus of 40% contrast (Gaussian-shaped and with a size similar to that of a Goldmann size III stimulus) is optimal; within the glaucoma cases, the amount of glaucomatous visual function loss correlated best with tracking performance at 160% contrast. Importantly, we also found that SONDA-EMP does not require significant learning, either in healthy individuals, nor in glaucoma patients^15^, or in patients with acquired brain injury (*Vrijling et al., submitted*). The next step is to build upon these findings by determining the screening performance of SONDA-EMP for glaucomatous visual function loss in a large sample of glaucoma cases and controls. In this way, we bring eye movement perimetry into the clinic.

Therefore, the aims of our current study were (1) to evaluate the screening performance of SONDA-EMP for detecting glaucomatous visual function loss and (2) to assess its user-friendliness. To examine the generalizability of the technique, we tested SONDA-EMP in two different implementations: SONDA-Eyelink (an experimental research setup^14,15^ and SONDA-Neon (a clinic-ready version). During testing, participants monocularly tracked a stimulus that moved continuously on a screen and made occasional jumps, requiring the participant to make both smooth pursuit and saccadic eye movements. These stimulus jumps were directed towards specific locations on the screen, in order to cover the test locations of the 24-2 grid of the Humphrey Field Analyzer (HFA; Carl Zeiss Meditec AG, Jena, Germany). We used tracking performance (continuously monitored agreement between stimulus and gaze position) as our outcome measure.^15^ In the present study, we determined the screening performance of SONDA-EMP in a cohort of 100 cases with early, moderate, or severe glaucoma, and 100 age- and sex-similar controls. For this, we built a Glaucoma Screening Score (GSS) using data from our previous study.^15^ Subsequently, we calculated, in the new data, the Area Under the ROC Curve (AUC) and determined the sensitivity at 95% specificity. The sensitivity was compared to predefined, stage-specific thresholds.^16^ User-friendliness was evaluated through a feedback questionnaire focusing on the ease, comprehensibility, and tiresomeness of the SONDA-EMP setups, and, for the cases, also of SAP.

## Methods

### Study population

In this study, we included 100 participants with glaucoma (cases) and 100 age- and sex-similar participants without glaucoma or other diseases causing VF defects (controls), aged between 42 and 92 years. Written informed consent was obtained after explanation of the nature and possible consequences of the study. As part of the consent procedure, participants were explicitly asked whether they consented to sharing their anonymised data with Reyedar. If a participant did not consent to the sharing, they could still participate in the study. The study was approved by the Medical Ethical Committee of the University Medical Center Groningen (NL84477.042.23). The study followed the tenets of the Declaration of Helsinki.

### Recruitment

Glaucoma patients who were scheduled for their routine clinical perimetry visit with SAP at the Ophthalmology Department of the University Medical Center Groningen (UMCG) were considered for inclusion. Patients were eligible to participate if they had a minimum of three consecutive reliable prior SAP assessments in which the Glaucoma Hemifield Test was reproducibly outside normal limits in at least one eye, fixation losses of ≤ 20%, and false positive (FP) responses ≤ 10%. The test pattern of these previous SAP assessments could be the HFA 24-2, 30-2, or 10-2, depending on the test pattern used clinically in the individual case. For all grids, the used thresholding strategy was SITA-Fast. Additional eligibility criteria included a best corrected visual acuity (BCVA) better than 0.3 logMAR in at least one eye, and absence of neurological disorders that could affect test performance. BCVA and neurological history were obtained from patients’ medical records; for neurological conditions, this information was verified using a short screening questionnaire. This questionnaire asked whether the patient was currently, or had previously been, under the care of a neurologist, and if so, for what reason. If all eligibility criteria were met, patients were invited to participate directly following their routine SAP evaluation. The reliability of this SAP assessment was subsequently evaluated. If the FP response exceeded the 10% threshold, inclusion was still allowed up to 15% FP, provided that (1) a clearly defined blind spot was visible, (2) no raw contrast threshold values exceeded 40 dB, and (3) the pattern deviation probability plot was not visibly darker than the total deviation probability plot. Similarly in the presence of increased fixation losses, fixation was considered acceptable if a well-defined blind spot was present or the technician had observed and reported stable fixation.^17,18^

If both eyes of a case met the inclusion criteria, one eye was selected for participation. Selection was random by default; however, prioritization was applied to ensure a representative distribution of glaucoma severity based on SAP mean deviation (MD) in decibels (dB) of the 24-2 or 30-2 test. To ensure clinically relevant evaluation of glaucoma screening sensitivity, we aimed for three approximately equal subgroups of glaucoma severity. This resulted in 36 cases with early (MD > –6 dB), 36 with moderate (–12 dB < MD ≤ –6 dB), and 28 with severe (MD < –12 dB) glaucoma. Additionally, efforts were made to balance the distribution of left and right eyes across glaucoma stages. Cases were allowed to have different types of glaucoma and concurrent other eye diseases, like myopia, as long as glaucoma was the presumed sole cause of the VF loss and loss of visual acuity, if any.

Controls were recruited via advertisement. Potential controls who responded to the advertisement were asked to fill out a questionnaire to screen for any known eye abnormalities, a positive family history of glaucoma, and neurological disorders that could affect test performance. During their visit, a short eye-health check was performed which included: an intraocular pressure (IOP) measurement (Ocular Response Analyzer G3 non-contact tonometer, Reichert Technologies, Inc., Depew, NY, USA), a refraction and BCVA measurement (Nidek ARK-1s. Nidek co., ltd, Gamagori, Japan), and a measurement of the peripapillary retinal nerve fiber layer thickness as assessed by spectral domain optical coherence tomography (OCT; Copernicus, Optopol Technologies, Zawierci, Poland). Exclusion criteria were any known eye abnormality, a positive family history of glaucoma, any known neurological disorders that could affect test performance, a BCVA worse than 0.3 logMAR, a cornea-compensated IOP (IOPcc) above 22 mmHg, or any temporally located red clock hour abnormality in the peripapillary retinal nerve fiber layer on the OCT. Controls where only one eye met the eligibility criteria were included, but only if both eyes were free of glaucomatous damage. The inclusion of controls was deliberately delayed relative to case inclusion, to enable control selection in order to reach a comparable age and sex distribution between cases and controls. In addition, the tested eye of the controls was selected to reflect the distribution of left and right eyes in the cases.

### SONDA-EMP setups

We implemented SONDA-EMP in two different setups: SONDA-Eyelink, our experimental research setup ^14,15^, and SONDA-Neon, a clinic-ready version (Reyedar B.V., Groningen, Netherlands). All participants underwent the assessments using both the SONDA-Eyelink and SONDA-Neon setups in a pseudorandomized order to minimize potential bias due to learning effects or fatigue. Both setups were placed in the same room. Testing was conducted in a dark room, with the only source of illumination being the monitor of the active setup. All tests were performed monocularly using optimal spherical equivalent refractive correction for the viewing distance.

### Stimulus and trajectories

The stimulus was a Gaussian blob with 0.22 degree standard deviation, corresponding to a diameter of a Goldmann size III stimulus (0.43 degrees), a commonly used stimulus size in VF testing. The stimulus was presented on a static gray background of 30 cd/m^2^ at a Weber contrast of 40% (both SONDA-EMP setups) and 160% (SONDA-Eyelink only).

The stimulus moved in saccadic pursuit mode. In this mode, the stimulus moves continuously, with pseudorandom variations in direction and velocity, and makes additional jumps to new VF locations. Spatially, the jumps were towards one of the 56 test locations of the HFA 24-2 grid (see Vrijling et al.^14^ for a description of the stimulus grid and for a detailed description of the generation of the saccadic pursuit trajectories). Each trajectory covered all 56 locations in three consecutive 40 s segments. A total of 15 unique trajectories were generated to minimize learning and predictive viewing behavior. Before the actual assessments, participants completed a separate practice trajectory at 640% contrast using the SONDA-Eyelink setup to become familiar with the task.

### SONDA-Eyelink setup

Participants were seated in front of a computer monitor at a viewing distance of 60 cm with their head placed on a chinrest and stabilized with a forehead rest to minimize head movements. Stimuli were presented on a 24.5-inch IPS monitor (OptixMag25R1X, Micro-Star International Co., Ltd., New Taipei City, Taiwan) with a framerate of 240 Hz. The monitor had a resolution of 1920 x 1080 pixels (49 x 29 degrees of visual angle at the viewing distance of 60 cm). The luminance of the monitor was calibrated as described previously.^14^ Stimulus display and gaze recording were controlled by the Psychtoolbox and Eyelink Toolbox extensions^19–22^ for MATLAB (The Mathworks, Inc., version 2021a). The PC for the stimulus presentation was an HP Elitedesk 800 G3 with a NVIDIA GeForce GTX 1080 graphics card, running Windows 10. An Eyelink Portable Duo eye-tracker (SR Research Ltd., Ottawa, Ontario, Canada), running software version 6.10.01, was used to measure participants’ eye movements. Monocular gaze data was acquired at a sampling frequency of 1000 Hz. The eye-tracker was (re)calibrated using the built-in nine-point calibration routine at the start of the experiment and after movement of the participant. Calibration accuracy was verified using the built-in validation routine and was accepted when the accuracy was defined ‘good’ by the tracker software (i.e., average error less than 0.5° and maximum error < 1.0°). If for at most one of the nine points the error was > 1.0° (usually one of the upper corners) and this could not be solved on repeated calibrations, the measurement was continued. The participant was excluded and replaced if a good calibration accuracy could not be achieved. With the described approach, one participant was excluded and replaced. A drift correction was performed prior to presenting each trajectory (see Methods section, subsection *Data collection*).

### SONDA-Neon setup

Participants were seated in front of a computer monitor at a viewing distance of 60 cm with their head placed on a chinrest to minimize head movements. Stimuli were presented on a 27-inch HP X27q QHD monitor with a framerate of 60 Hz. The monitor had a resolution of 1920 x 1080 pixels (53 x 31 degrees of visual angle at the viewing distance of 60 cm), and was calibrated according to the procedure described in Vrijling et al.^14^. Stimulus display and gaze recording were controlled by PsychoPy^23^ and Pupil-labs Neon companion app on a OnePlus 10 Pro. The PC for the stimulus presentation was an Intel NUC12WSKi5. Monocular gaze data of the participants were acquired at a sampling frequency of 200 Hz using the Pupil-labs Neon eyetracker.^24^ The eyetracker employs a deep-learning-based pipeline that does not require prior user-specific calibration and has a median per-subject accuracy of 1.8°. The stimulus trajectories were generated at 60 Hz to match the screen refresh rate. A scene camera mounted on the eye tracker recorded a view of the environment at a framerate of 10 Hz, detecting the computer display in the scene video via edge detection. This information was used to compensate for residual head movements of the participant by interpolating the original trajectory with the adjusted computer display position. The head-movement-corrected stimulus trajectory was then upsampled to 200 Hz to match the gaze sampling frequency.

### Data collection

Cases performed their study related assessments following their routine clinical perimetry visit. Each participant started with the practice trajectory at the SONDA-Eyelink setup, followed by the main assessment on either the SONDA-Neon or the SONDA-Eyelink setup, presented in a pseudorandomized order across the participants to balance effects related to learning and fatigue, as well as carry-over effects on the user-friendliness, if any. For the SONDA-Eyelink setup, the order of the contrast levels was pseudo-randomized such that the contrast level order was balanced within the groups (cases/controls). Participants were instructed to follow the moving stimulus with their gaze to the best of their abilities, without moving their head. Overall, the SONDA-Eyelink assessment lasted about 8 minutes, including eye-tracker calibration and testing at two contrast levels, while the SONDA-Neon assessment took about 5 minutes (no calibration needed, and only one contrast level).

### Evaluation of user-friendliness

To qualitatively assess the user-friendliness of the SONDA-EMP setups, participants were asked to fill out a short feedback questionnaire after each assessment. The questionnaire consisted of three questions evaluating 1) the ease of the task, 2) the comprehensibility (clarity of the instructions), and 3) the tiresomeness of the test. Each item was rated on a scale from 1 to 10, with higher scores indicating a more positive experience. Participants completed the questionnaire after each assessment with the SONDA-Eyelink and SONDA-Neon. In addition, cases were also asked to complete the same questionnaire following their clinical SAP assessment.

### Data analyses

#### Pre-processing of eye movement data

Eye gaze positions were recorded in horizontal and vertical screen coordinates (pixels) and analyzed separately. Before calculation of tracking performance, any unreliable signal was filtered out of the raw gaze data. Unreliable signal was defined as velocity spikes (samples with a velocity larger than 750 deg/s), which are not likely to be caused by actual eye movements^25,26^ and position plateaus (multiple samples with zero velocity). The position plateaus were a consequence of how the gaze data was stored; gaze positions were stored in matrices at every frame to ensure the stimulus and gaze vectors were time linked. If tracking failed during stimulus presentation, those gaze positions were replaced by the last known gaze position.

The unreliable samples and an additional 0.05 seconds of data preceding and following each unreliable signal period (to remove any additional noise surrounding blinks) were filtered out by setting them to NaN. If more than half of the 40 s segment (in horizontal or vertical direction) was set to NaN after this step, it was discarded by setting the output parameters to NaN. Using this criterion, one 40 s segment (0.06% of the total number of trajectories) was discarded; this was one 40 s segment from a control participant where the eye-tracking failed for about 26 s.

In the SONDA-Neon setup, blinks manifest as narrow positive or negative peaks in the horizontal or vertical gaze position (without plateaus). These are detected by identifying pairs of velocity spikes of opposite sign occurring within 150 ms of each other. Each detected blink is replaced by a linear interpolation of eye position between the onset and offset of the blink. Using the same criterion as for the SONDA-Eyelink setup, no data segments were excluded due to excessive unreliable data.

### Definition of tracking performance

First, we calculated the cosine similarity (normalized inner product) between the stimulus and gaze vector (Eq. 1):

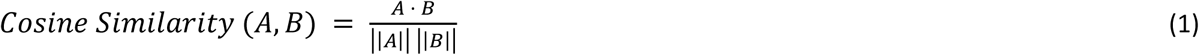

where A is the vector of stimulus positions and B the vector of gaze positions, ||A|| the Euclidean norm of vector A, and ||B|| the Euclidean norm of vector B. Theoretically, the cosine similarity is bound between −1 and 1. However, in practice, it ranges between zero — denoting poor agreement between the vectors (i.e., bad performance) — and one — denoting perfect agreement (i.e., excellent performance). Before calculating the cosine similarity, vectors A and B were both mean centered. Obviously, there is a (physiological) delay between stimulus and response. Therefore, we calculated a series of ‘time-shifted’ cosine similarities, with a time shift in the range of −5 and +5 seconds, in steps of 0.0042 seconds (the duration of 1 frame at 240 Hz). Next, we identified the first positive peak in the cosine similarity versus the time shift curve (‘cosine similarity function’) after 0 seconds (because of causality) using the Matlab function *findpeaks*. Tracking performance was defined as the cosine similarity corresponding to this peak. If no clear peak could be identified, the maximum cosine similarity at time shifts between 0 and +3 seconds was used. For each saccadic pursuit trajectory, the cosine similarity peaks from the three 40 s segments were averaged to obtain the mean tracking performance for the concerning trajectory. See Vrijling et al.^15^, Figure 1 for an example of a cosine similarity function of a control and a glaucoma case. Horizontal and vertical tracking performance were averaged prior to the statistical analyses. In our previous work, we demonstrated that the effects of stimulus contrast, age, and stage on horizontal and vertical tracking performance were highly similar^14,15^; *Vrijling et al. submitted*), supporting the use of the average.

**Figure 1.**
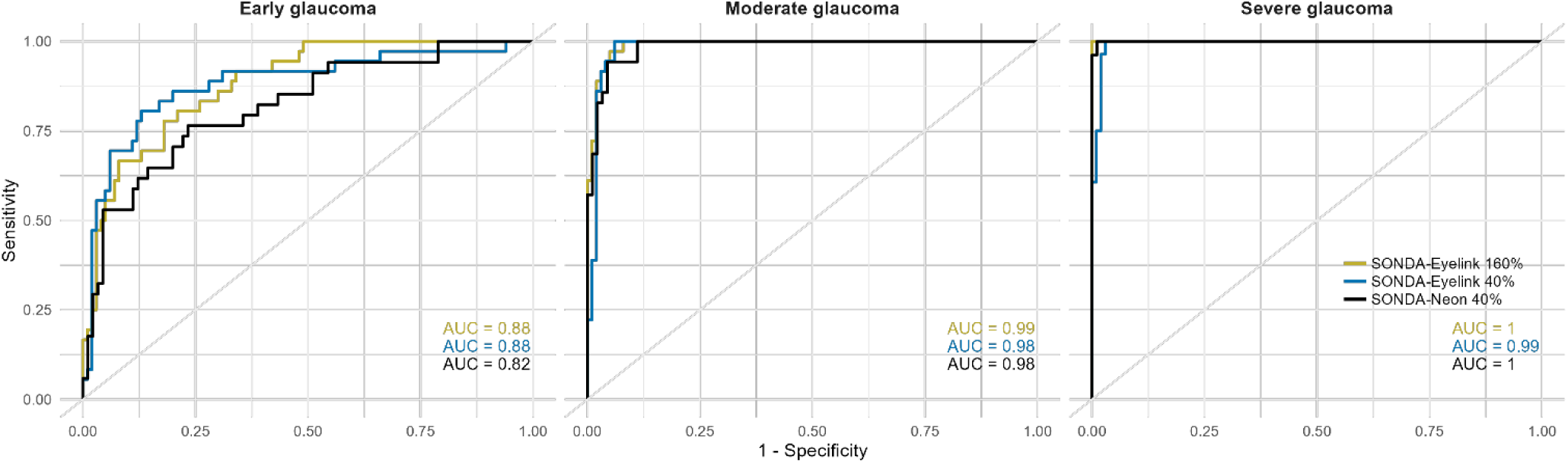
Receiver Operator Characteristic curves per glaucoma stage for the SONDA-Eyelink setup at 40% contrast (blue curves) and 160% contrast (yellow curves) and the SONDA-Neon setup at 40% contrast (black curves). The Area under the curve (AUC) values for each setup are shown in corresponding colors.

### Statistical analyses

All statistical analyses were conducted in R (version 4.2.2) using R Studio (version 1.3.1093). The study population was described using nonparametric descriptive statistics (median with interquartile range [IQR]).

Univariable comparisons between cases and controls were performed using the Mann–Whitney U test for continuous variables (age, spherical equivalent, visual acuity), and the Pearson’s Chi-squared test for categorical variables (sex and tested eye). For comparing the glaucoma subgroups, one-way Anova was used for continuous variables (age, spherical equivalent, visual acuity, and SAP mean deviation [MD]) in decibels), and Pearson’s Chi-squared test for proportions (sex and tested eye).

### Glaucoma Screening Score (GSS)

To validate the screening performance of SONDA-EMP tracking performance, we calculated a Glaucoma Screening Score (GSS) based on the tracking performance and age of a participant using the following equations:

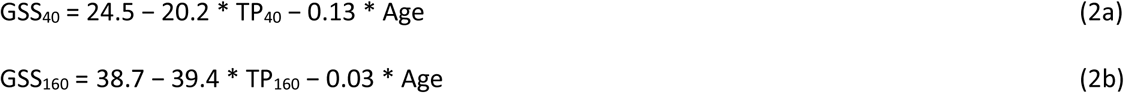

where TP_40_ and TP_160_ are the tracking performances at 40% and 160% contrast. Since no previously collected case-control dataset was available for the SONDA-Neon setup, we considered the GSS as specified in Eq (2a) to be a generic GSS, not bound to a specific setup.

The parameters in these equations were obtained by conducting a logistic regression analysis on previously acquired data using 36 cases and 36 controls.^15^ In this analysis, case-control was used as the dependent variable, while TP (40% or 160% contrast) was the independent variable. Age was included as a covariate. Note that this pre-existing dataset has been acquired with the same SONDA-Eyelink setup as used in this study.

### Screening performance of SONDA-EMP for detecting glaucomatous visual function loss

We calculated the GSS of each participant in the current study and performed receiver operator characteristic (ROC) analyses, separately for SONDA-Eyelink at 40% contrast, SONDA-Eyelink at 160% contrast, and SONDA-Neon at 40% contrast. We calculated the Area Under the ROC Curve (AUC) and determined the sensitivity at 95% specificity. This was done separately for each glaucoma stage. The sensitivity was compared to pre-defined, stage-specific thresholds, being 50% for early glaucoma, 90% for moderate glaucoma, and 100% for severe glaucoma.^16^ We defined a good screening performance as a sensitivity at or above these thresholds, at 95% specificity.

### User-friendliness of SONDA-EMP setups

We compared the scores for each questionnaire item between cases and controls for both SONDA-EMP setups separately using the Wilcoxon signed-rank test. Subsequently, we compared the scores for the glaucoma cases across both SONDA-EMP setups and SAP using Friedman’s test. Where Friedman’s test was significant, post-hoc pairwise comparisons were performed using the Wilcoxon signed-rank test. To control for multiple comparisons within each item, the *P* values from the Wilcoxon tests were adjusted using Bonferroni correction. The effect of glaucoma stage on the scoring of the cases was evaluated with Kruskal-Wallis tests.

## Results

Table 1 shows the general characteristics of the study population. Cases and controls did not differ significantly regarding age, sex, or eye tested. Compared to controls, cases had a reduced visual acuity (higher logMAR value) and were on average more myopic. The median VF MD (HFA) of the included eye of the glaucoma cases was approximately −8 dB.

**Table 1.**
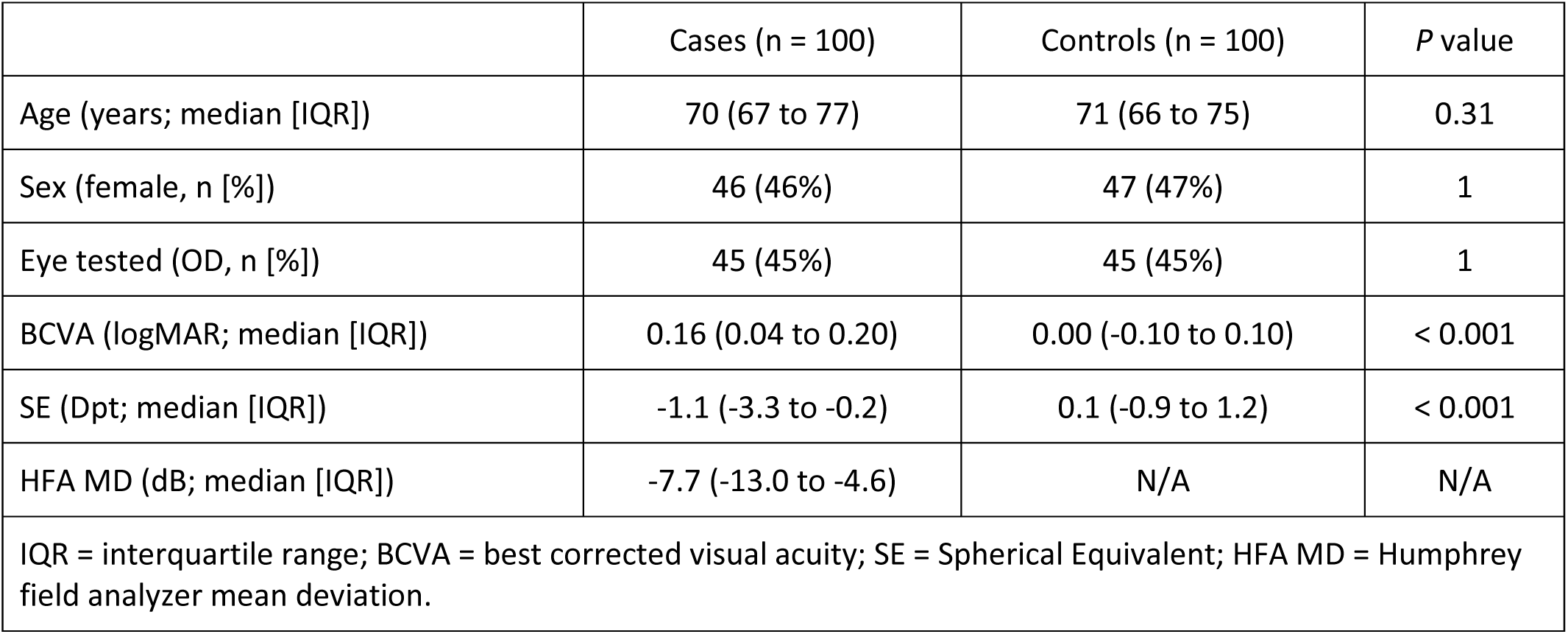
Characteristics of the study population.

Table 2 shows the characteristics of the glaucoma cases, per glaucoma stage. There were no differences regarding age, sex, eye tested, BCVA, or refractive error (SE) between the stages.

**Table 2.** Characteristics of the glaucoma cases (n = 100).

|  | Early (n = 36) | Moderate (n = 36) | Severe (n = 28) | P value |
| --- | --- | --- | --- | --- |
| Age (years; median [IQR]) | 71 (68 to 77) | 70 (64 to 75) | 72 (67 to 80) | 0.18 |
| Sex (female; n [%]) | 19 (53%) | 16 (44%) | 11 (39%) | 0.75 |
| Eye tested (OD; n [%]) | 17 (47%) | 16 (44%) | 12 (43%) | 0.99 |
| BCVA (logMAR; median [IQR]) | 0.10 (0.04 to 0.20) | 0.20 (0.07 to 0.21) | 0.15 (0.05 to 0.20) | 0.28 |
| SE (Dpt; median [IQR]) | -1.0 (-3.3 to 0.0) | -1.1 (-3.0 to -0.2) | -1.3 (-3.8 to -0.4) | 0.86 |
| HFA MD (dB; median [IQR]) | -3.4 (-4.8 to -2.2) | -8.1 (-10.0 to -7.3) | -18.0 (-20.7 to -14.5) | N/A |

IQR = interquartile range; BCVA = best corrected visual acuity; SE = Spherical Equivalent; HFA MD = Humphrey field analyzer mean deviation.

Figure 1 shows the ROC curves for the GSS per glaucoma stage and per SONDA-EMP setup.

Table 3 summarizes the screening performance of the SONDA-Eyelink setup at 40% and 160% contrast. For all glaucoma stages, and at both contrast levels, the screening performance met the predefined thresholds for good screening performance.

**Table 3.** Screening performance for all glaucoma cases together and separately for each glaucoma stage and contrast level for the SONDA-Eyelink setup.

| Contrast | Glaucoma Stage | AUC (95% CI) | Number of true positives | Number of false negatives | Sensitivity at 95% specificity (95% CI) |
| --- | --- | --- | --- | --- | --- |
| 40% | All stages | 0.95 (0.92 – 0.98) | 83 | 17 | 0.83 (0.75 – 0.89) |
|  | Early | 0.88 (0.81 – 0.96) | 21 | 15 | 0.58 (0.42 – 0.73) |
|  | Moderate | 0.98 (0.95 – 1.00) | 34 | 2 | 0.94 (0.83 – 0.99) |
|  | Severe | 0.99 (0.96 – 1.00) | 28 | 0 | 1.00 (0.90 – 1.00) |
| 160% | All stages | 0.95 (0.92 – 0.98) | 83 | 17 | 0.83 (0.75 – 0.89) |
|  | Early | 0.87 (0.80 – 0.95) | 20 | 16 | 0.56 (0.39 – 0.71) |
|  | Moderate | 0.99 (0.96 – 1.00) | 35 | 1 | 0.97 (0.87 – 1.00) |
|  | Severe | 1.00 (0.99 – 1.00) | 28 | 0 | 1.00 (0.90 – 1.00) |
| AUC = area under the curve; CI = confidence interval. |  |  |  |  |  |

The SONDA-Neon dataset initially included 100 controls and 98 glaucoma cases. Two glaucoma cases (one with early glaucoma and one with severe glaucoma) did not consent to share their data with Reyedar, who extracted the data out of the device with proprietary software. After testing, thirteen participants (ten controls and three glaucoma cases - one per glaucoma stage) were excluded due to initial data storage failures (8) and eyetracker failures (5). As a result, data from 90 controls and 95 glaucoma cases were included in the final analysis.

Table 4 summarizes the screening performance of the SONDA-Neon setup. For all glaucoma stages, and at both contrast levels, the screening performance met the predefined thresholds for good screening performance.

**Table 4.** Screening performance of the GSS for all glaucoma cases and separately for each glaucoma stage for the SONDA-Neon setup.

| Glaucoma stage | AUC (95% CI) | Number of true positives | Number of false negatives | Sensitivity at 95% specificity (95% CI) |
| --- | --- | --- | --- | --- |
| All stages | 0.93 (0.89 - 0.96) | 77 | 18 | 0.81 (0.72 – 0.88) |
| Early | 0.82 (0.72 - 0.91) | 18 | 16 | 0.53 (0.36 – 0.69) |
| Moderate | 0.98 (0.95 - 1.00) | 33 | 2 | 0.94 (0.82 – 0.99) |
| Severe | 1.00 (0.99 - 1.00) | 26 | 0 | 1.00 (0.89 – 1.00) |
| AUC = area under the curve; CI = confidence interval. |  |  |  |  |

### User-friendliness

Table 5 shows, on a scale between 1 and 10 with 10 meaning easier, more comprehensive, and less tiresome, that both cases and controls gave favorable scores to both SONDA-EMP setups, irrespective of the specific setup used. Controls rated both SONDA-EMP setups as significantly easier to perform and more comprehensible than the glaucoma cases did. There were no statistically significant differences in reported tiresomeness of the test between cases and controls.

**Table 5.** User-friendliness scores (median [IQR]) for SONDA-Eyelink and SONDA-Neon setups for glaucoma cases and controls.

|  | Cases (n = 100) | Controls (n = 100) | P value |
| --- | --- | --- | --- |
| Ease of test (median (IQR)) |  |  |  |
| SONDA-Eyelink | 8.3 (7.0 – 9.0) | 10 (9.0 – 10) | < 0.001 |
| SONDA-Neon | 9.0 (8.0 – 10) | 10 (9.0 – 10) | < 0.001 |
| Comprehensibility of test (median (IQR)) |  |  |  |
| SONDA-Eyelink | 10 (9.0 – 10) | 10 (9.5 – 10) | 0.004 |
| SONDA-Neon | 10 (9.0 – 10) | 10 (10 – 10) | < 0.001 |
| Tiresomeness of test (median (IQR)) |  |  |  |
| SONDA-Eyelink | 8.0 (7.0 – 9.0) | 8.0 (8.0 – 10) | 0.07 |
| SONDA-Neon | 9.0 (8.0 – 9.3) | 9.0 (8.0 – 10) | 0.06 |

Table 6 shows that the ease and tiresomeness are scored differently between the three setups (Eyelink, Neon, and SAP). Comprehensibility scores were high across all setups, with no significant differences.

**Table 6.** User-friendliness scores (median [IQR]) for SONDA-Eyelink, SONDA-Neon, and SAP among glaucoma cases.

|  | SONDA-Eyelink | SONDA-Neon | SAP | *P* value |
| --- | --- | --- | --- | --- |
| Ease of test (median (IQR)) | 8.3 (7.0 – 9.0) | 9.0 (8.0 – 10) | 7.3 (6.0 – 9.0) | < 0.001 |
| Comprehensibility of test (median (IQR)) | 10 (9.0 – 10) | 10 (9.0 – 10) | 10 (8.0 – 10) | 0.34 |
| Tiresomeness of test (median (IQR)) | 8.0 (7.0 – 9.0) | 9.0 (8.0 – 9.3) | 6.0 (4.0 – 8.0) | < 0.001 |

Post-hoc Tukey tests showed that SONDA-Neon was rated significantly easier than SONDA-Eyelink (*P* = 0.004) and SAP (*P* < 0.001). SONDA-Eyelink was also easier than SAP (*P* = 0.01). SONDA-Neon was considered significantly less tiring than both SONDA-Eyelink and SAP (*P* < 0.001), with SONDA-Eyelink also rated less tiring than SAP (*P* < 0.001). No statistically significant differences were found across early, moderate, and severe glaucoma cases in ease, comprehensibility, or tiresomeness for any of the three test setups (see Table 7).

**Table 7.** Comparison of user-friendliness scores (median [IQR]) per glaucoma stage for SONDA-Eyelink, SONDA-Neon, and SAP.

|  | Setup | Early | Moderate | Severe | *P* value |
| --- | --- | --- | --- | --- | --- |
| Ease of test (median (IQR)) | SONDA-Eyelink | 9.0 (7.3 – 10) | 8.0 (7.0 – 9.0) | 8.0 (6.0 – 9.0) | 0.15 |
|  | SONDA-Neon | 9.0 (8.0 – 10) | 9.0 (8.0 – 9.0) | 8.0 (7.0 – 9.0) | 0.15 |
|  | SAP | 8.0 (6.0 – 8.8) | 7.0 (6.0 – 8.5) | 7.0 (6.0 – 8.5) | 0.98 |
| Comprehensibility of test<br>(median (IQR)) | SONDA-Eyelink | 9.0 (8.3 – 10) | 10 (9.0 – 10) | 9.0 (9.0 – 10) | 0.10 |
|  | SONDA-Neon | 9.0 (9.0 – 10) | 10 (9.0 – 10) | 10 (9.0 – 10) | 0.09 |
|  | SAP | 9.5 (8.0 – 10) | 10 (9.0 – 10) | 9.0 (8.0 – 10) | 0.57 |
| Tiresomeness of test<br>(median (IQR)) | SONDA-Eyelink | 9.0 (7.3 – 9.4) | 8.0 (6.0 – 9.0) | 8.0 (6.5 – 9.0) | 0.10 |
|  | SONDA-Neon | 9.0 (8.0 – 10) | 8.0 (8.0 – 10) | 8.0 (8.0 – 9.0) | 0.09 |
|  | SAP | 7.0 (6.0 – 8.0) | 6.0 (3.5 – 7.5) | 7.0 (3.5 – 8.0) | 0.11 |

## Discussion

The main finding of this study is that SONDA-EMP is an effective method for detecting glaucomatous visual function loss. Both SONDA-Eyelink and SONDA-Neon met the predefined sensitivity thresholds at 95% specificity of 50, 90, and 100% for early, moderate, and severe glaucoma, respectively.^16^ Cases and controls considered SONDA-EMP as easy, comprehensible, and less tiresome. Cases rated SONDA-EMP consistently easier and less tiring compared to SAP.

Our finding that eye movement perimetry (EMP) has a good screening performance across all stages of glaucoma is in agreement with previous research using saccadic reaction times (SRT) for glaucoma detection.^6,8^ Kadavath Meethal et al.^8^ reported an overall AUC of 0.95, with a sensitivity of 87.7% at 96.2% specificity. Their study included 104 controls (58 males) and 73 glaucoma cases (54 males) stratified into three groups: 33 with early glaucoma, 23 with moderate glaucoma, and 17 with severe glaucoma. Groups had a similar age distribution (mean ± SD; 52 ± 13 years and 49 ± 14 years for glaucoma cases and controls, respectively; *P* = 0.15) but differed regarding sex, with more males in the glaucoma group. Mazumdar et al.^6^ reported a sensitivity of 100% at a specificity of 93% to 96.4% (across two independent glaucoma specialists). Their study included 28 controls and 24 glaucoma cases, 4 with early glaucoma, 11 with moderate glaucoma, and 9 with severe glaucoma. The mean VF Mean Deviation (MD) for the glaucoma groups was −14.4 (SD 9.4) dB, indicating more advanced disease compared to our sample (−7.7 dB). Group differences were observed in age (53 ± 13 years and 42 ± 14 years for the glaucoma cases and controls, respectively), they adjusted their analysis for this difference by deriving age-specific normative SRT thresholds based on a database of 95 controls stratified into five age bins^11^, and sex, where there were more males in the glaucoma group. In our study, we observed an overall AUC ranging from 0.93 to 0.95, with a sensitivity of 0.81 to 0.83 at 95% specificity, depending on stimulus contrast and setup used.

These results are comparable to those reported by Kadavath Meethal et al.^8^, who included patients across a similar range of glaucoma severities. The findings by Mazumdar et al.^6^ are comparable to our results for the subgroup of patients with severe glaucoma; however, their higher overall sensitivity may in part reflect the inclusion of more severe cases. Overall, SONDA-EMP achieves a screening performance comparable to previously reported EMP approaches. However, our method is performed continuously, without requiring participants to refixate between stimuli. This contrasts with earlier EMP tests^4–12^, which do require repeated refixations. As a result, SONDA-EMP allows for a more efficient and intuitive testing procedure, potentially reducing test duration and participant burden while maintaining screening performance.

Participants reported that SONDA-EMP was easy to perform, well-understood, and less tiring than SAP, with the SONDA-Neon setup rated most favorably overall (the difference between SONDA-Eyelink and SONDA-Neon in tiresomeness but not easiness could be explained by the two contrast levels tested with SONDA-Eyelink in the current research setting). These results align with previous research on EMP usability in super elderly.^27^

This study has strengths and limitations. The analyses were based on the tracking performance, one of various spatiotemporal properties (STPs) that can potentially be used as a metric for EMP. To verify the validity of this choice, we explored whether adding other STPs could provide additional information. Adding other STPs did not improve the screening performance. The GSS that we used in this study, was based on previously collected data, to enable evaluation of SONDA-EMP screening performance in independently collected data. The previously collected data were collected with the SONDA-Eyelink setup. As such, the GSS might be biased towards this setup. We did not observe a clear difference in screening performance between both setups; the apparent difference in sensitivity for early glaucoma was not statistically significant (*P* = 0.83). Hence, either the GSS is not setup-specific or the SONDA-Neon could perform even better if it had its own GSS. To explore this, we estimated the screening performance of SONDA-Neon with a stratified 10-fold cross-validation resampling technique^28^, without using the SONDA-Eyelink-based GSS. A very similar screening performance was found (see Supplementary material). This supports the generalizability of the SONDA-EMP technique, which is apparently robust to differences in, for example, hardware, sample rate, calibration procedure, and measurement noise. A subset of the participants in this study (25 controls and 6 glaucoma cases; 1 case with early glaucoma, 3 with moderate glaucoma and 2 with severe glaucoma) also participated in our previous study.^15^ They responded to both calls for participation, which was initially overlooked. Although these participants were re-tested in the current study, their measurements were conducted independently and often by different technicians/researchers, and there was an interval of approximately 2 years between both studies. Their inclusion means that the two datasets were largely, but not completely independent. As such, this may have led to a slight overestimation of the screening performance of SONDA-Eyelink. To address this, we repeated the ROC analysis described in the Methods section (subsection *Screening performance of SONDA-EMP for detecting glaucomatous visual function loss*) after excluding all previously tested participants (resulting in a reduced sample of 169 participants; 94 cases and 75 controls). The results were highly consistent (see Table S.1 supplementary material), indicating that the unintentional overlap has not influenced our results and conclusions. Initially, the assessment with the SONDA-Neon setup gave some technical difficulties, resulting in the loss of data of 13 participants. These technical problems were solved, no further data loss occurred during the study, and no further impact on future data collection is expected.

Our results show that SONDA-EMP is ready for clinical implementation, particularly for glaucoma screening. Both SONDA-EMP setups met predefined screening performance thresholds across glaucoma stages. While the sensitivity for detecting early glaucoma may seem modest (53–58% at 95% specificity), this level is on par with that of other perimetric screening tests, such as Frequency Doubling Technology (FDT).^29–31^ For population-based screening, detecting half of the early glaucoma cases and virtually all later stages is considered acceptable. A higher sensitivity would imply a lower specificity, whereas a high specificity is essential to reduce false positives and ensure cost-effectiveness. This requirement makes detection of early glaucoma inherently challenging. In addition to its screening performance, an advantage of the SONDA-EMP technique is that its test duration remains consistent across all stages of glaucoma. Unlike conventional threshold-based perimetric approaches, such as SAP, where more advanced visual field loss typically results in prolonged testing times, SONDA-EMP’s fixed test duration ensures a time-efficient assessment irrespective of glaucoma stage. This characteristic may be especially beneficial for clinical populations with limited sustained attention, fatigue issues, or cognitive impairment, and likely contributed to the favorable subjective evaluations observed even in participants with severe visual function loss. Future studies should focus on investigating the screening performance of SONDA-EMP in diverse populations, such as children or those with acquired brain injury.

In this study, we evaluated screening performance at two contrast levels for the SONDA-Eyelink setup: 40% and 160%. Our current results show that 40% and 160% contrast (in combination with stimulus size III) perform equally well in detecting early glaucoma, where 160% contrast is even slightly better for cases with moderate glaucoma. Our earlier work showed that visual function loss (in terms of SAP Mean Sensitivity) could be estimated best from the tracking performance for a stimulus contrast of 160%, and may therefore be better suited for disease monitoring.^15^ These findings together suggest that 160% contrast could serve a dual role — both in screening and in monitoring previously diagnosed glaucoma cases. A longitudinal study is pivotal to investigate whether 160% contrast is suitable for monitoring glaucoma progression.

### Conclusion

There is a need for fast, intuitive, and accessible methods to screen for visual function loss in glaucoma. SONDA-EMP meets this need. Beyond screening and staging, further steps could focus on validating its use for monitoring of previously diagnosed glaucoma cases, establishing usability in and age correction for a wider age range, and validating SONDA-EMP in other pathologies, such as acquired brain injury, to evaluate its applicability beyond glaucoma.

## Data Availability

The data are available at the DataverseNL repository via https://doi.org/10.34894/RGVXPY, while materials (experimental code) are available at request from the corresponding author. The experiments were not pre-registered.

## Acknowledgements

The authors thank Kim Westra for the recruitment of glaucoma patients. The authors also thank Dorien Drent, Jip Konings, Luisa Bongaerts, Olga Leciejewska, and Erwin van der Staaij for their help with the measurements of participants.

## Conflict of interest

RJR is listed as inventor on the patent WO2021096361A1 (Grillini, A., Hernández-García, A., Renken, R. J. 2019; Method, system, and computer program product for mapping a visual field) on which the method used in this manuscript is partially based.

The UMCG conducted the experiments and performed the statistical analyses. Reyedar B.V. Groningen, the Netherlands calculated the tracking performance from the SONDA-Neon data.

