## Supplementary material for "Screening for glaucoma with a novel eye movement perimetry technique based on continuous visual stimulus tracking"

*Cross-validation procedure*

The glaucoma screening score (GSS) used in this study was derived from a previously collected dataset. This reference dataset was obtained with the SONDA-Eyelink setup, and as such, the GSS could in principle be biased toward that setup. To validate its applicability to the present SONDA-Neon dataset, we performed a stratified 10-fold cross-validation[1] using SONDA-Neon data only. For this, the control participants were equally distributed over the 10 folds. Glaucoma cases were divided per disease stage (early, moderate, severe), and per stage, they were approximately equally distributed over the 10 folds. This ensured that each fold contained a representative distribution of the glaucoma stages and an equal ratio of cases and controls.

For each of the 10 iterations, nine folds were used to derive the GSS, and the remaining single fold was used to evaluate the screening performance. For deriving the GSS, a logistic regression model with case versus control as the dependent variable, the tracking performance as independent variable, and age as a covariate was fitted to the data in the combined nine folds. For each participant, a GSS was calculated from the resulting model coefficients. The classification threshold was defined as the 95th percentile of the GSS distribution among the controls in the nine folds, corresponding to 95% specificity. This threshold was then applied to the remaining single fold to calculate the sensitivity, for each glaucoma stage separately. This was repeated ten times, such that each fold was used exactly once for evaluation, resulting in mean sensitivity values with 95% confidence intervals (CIs). The resulting mean sensitivities (95% CI) at 95% specificity were: early glaucoma 0.60 (0.41–0.79), moderate glaucoma 0.92 (0.82–1.00), and severe glaucoma 1.00 (1.00–1.00). This indicates good screening performance when compared to the pre-defined, stage-specific thresholds, being 50% for early glaucoma, 90% for moderate glaucoma, and 100% for severe glaucoma.[2]

*Limited influence of partial participant overlap with previous study*

Table S.1 summarizes the screening performance of the SONDA-Eyelink setup at 40% and 160% contrast, with all participants included and after exclusion of previously tested participants.[3]

**Table S.1**. Screening performance per glaucoma stage and contrast level for the SONDA-Eyelink setup, with all data included and after exclusion of previously tested participants (shown in rows marked "- subgroup")

| Contrast | Glaucoma stage | AUC (95% CI) | Number of true positives | Number of false negatives | Sensitivity at 95% specificity (95% CI) |
|---|---|---|---|---|---|
| 40% | Early | 0.88 (0.81 – 0.96) | 21 | 15 | 0.58 (0.42 – 0.73) |
| | Early - subgroup | 0.87 (0.79 – 0.95) | 20 | 15 | 0.57 (0.41 – 0.73) |
| | Moderate | 0.98 (0.95 – 1.00) | 34 | 2 | 0.94 (0.82 – 0.98) |
| | Moderate - subgroup | 0.98 (0.94 – 1.00) | 32 | 1 | 0.97 (0.86 –1.00) |
| | Severe | 0.99 (0.96 – 1.00) | 28 | 0 | 1.00 (0.90 – 1.00) |
| | Severe - subgroup | 0.99 (0.96 – 1.00) | 26 | 0 | 1.00 (0.89 – 1.00) |
| 160% | Early | 0.87 (0.80 – 0.95) | 20 | 16 | 0.56 (0.39 – 0.71) |
| | Early - subgroup | 0.84 (0.75 – 0.93) | 18 | 17 | 0.51 (0.35 – 0.68) |
| | Moderate | 0.99 (0.96 – 1.00) | 35 | 1 | 0.97 (0.87 – 1.00) |
| | Moderate - subgroup | 0.98 (0.95 – 1.00) | 31 | 2 | 0.94 (0.81 – 0.99) |
| | Severe | 1.00 (0.99 – 1.00) | 28 | 0 | 1.00 (0.90 – 1.00) |
| | Severe - subgroup | 1.00 (0.99 – 1.00) | 26 | 0 | 1.00 (0.89 – 1.00) |
| AUC = area under the curve; CI = confidence interval. | | | | | |

*References*


1. Kohavi R. A study of cross-validation and bootstrap for accuracy estimation and model selection. In: *Proceedings of the 14th International Joint Conference on Artificial Intelligence - Volume 2*. IJCAI'95. Morgan Kaufmann Publishers Inc.; 1995:1137-1143.

2. Stoutenbeek R. *Population Based Glaucoma Screening*. University Library Groningen; 2010.

3. Vrijling ACL, De Boer MJ, Renken RJ, et al. Detecting and Quantifying Glaucomatous Visual Function Loss With Continuous Visual Stimulus Tracking: A Case-Control Study. *Trans Vis Sci Tech*. 2025;14(2):3. doi:10.1167/tvst.14.2.3.
